# A comparison of fMRI presurgical mapping techniques with intraoperative brain mapping-based validation

**DOI:** 10.1101/2023.10.17.23297054

**Authors:** Ahmed M. Radwan, Louise Emsell, Kristof Vansteelandt, Evy Cleeren, Ronald Peeters, Steven De Vleeschouwer, Tom Theys, Patrick Dupont, Stefan Sunaert

**Affiliations:** KU Leuven, Department of Imaging and pathology, Translational MRI, Leuven, Belgium; KU Leuven, Leuven Brain Institute (LBI), Department of Neurosciences, Leuven, Belgium; KU Leuven, Department of Neurosciences, Neuropsychiatry, Leuven, Belgium; KU Leuven, Department of Geriatric Psychiatry, University Psychiatric Center (UPC), Leuven, Belgium; UZ Leuven, Department of Neurology, Leuven, Belgium; UZ Leuven, Department of Radiology, Leuven, Belgium; KU Leuven, Department of Neurosciences, Research Group Experimental Neurosurgery and Neuroanatomy, Leuven, Belgium; UZ Leuven, Department of Neurosurgery, Leuven, Belgium; KU Leuven, Laboratory for Cognitive Neurology, Department of Neurosciences, Leuven, Belgium

## Abstract

**Introduction:** Resting-state functional MRI (rs-fMRI) could enable preoperative risk assessment and intraoperative guidance for patients who cannot undergo task-based fMRI (tbfMRI). We investigated differences in accuracy between tbfMRI and rsfMRI acquired with single (sTE) at voxel size of 2mm and 3mm, and multi-echo (mTE) scans using intraoperative mapping with direct electrical stimulations (DES) as the ground truth.

**Material and methods:** Functional sensory-motor mapping results of hands and feet were spatially compared relative to positive (pDES, functional effect) and negative (nDES, no functional effect) coordinates in 16 preoperative patients. General linear model analysis was used for tbfMRI, and seed-based analysis (SBA) for rsfMRI. Minimum Euclidean distances between fMRI and DES were calculated and compared between fMRI methods. Receiver-operating characteristic (ROC) curves were used to compare accuracy and determine distance cutoffs for fMRI agreement with DES, and binary agreement rates were compared at different cutoffs. Two-part mixed-effects linear models were used to compare fMRI methods while accounting for unequal inter-subject DES repetition.

**Results:** Only minor differences were found between fMRI methods in unthresholded distances (mean differences ∼ 2 mm). ROCs and binary agreement measures showed comparable accuracy for tbfMRI and sTE-rsfMRI 2mm, but mildly worse for sTE-rsfMRI 3mm and mTE-rsfMRI. However, differences in relative accuracy between sTE- and mTE-rsfMRI were minor when the same distance cutoff was applied to all methods. This was also reflected on comparing rates of binary agreement and confirmed by the two-part linear models, which showed no significant differences between fMRI methods and a significant effect of DES response.

**Conclusion:** The similar accuracy for SBA rsfMRI functional sensory-motor mapping compared to tbfMRI for the hands and feet, indicate that rsfMRI may be suitable for presurgical mapping. The differences in relative accuracy between sTE- and mTE rsfMRI warrant further investigation in a larger sample.

## Introduction

Blood oxygen-level dependent (BOLD) task-based functional magnetic resonance imaging (tbfMRI) is routinely used in clinical practice for presurgical brain mapping. TbfMRI is typically used for sensory-motor and language mapping and for estimating hemispheric language dominance (Lehéricy et al., 2000; Stippich et al., 2007; Sunaert, 2006; Wengenroth et al., 2011). Functional brain mapping with tbfMRI has been previously validated against intraoperative direct electrical stimulation (DES) (Fandino et al., 1999; Jack et al., 1994; Lehéricy et al., 2000; Xie et al., 2008), which is the gold-standard method for functional brain mapping. The use of fMRI for presurgical brain mapping was also recently shown to be associated with decreased morbidity and mortality in brain tumor patients undergoing surgical resection (Vysotski et al., 2018). BOLD tbfMRI can be used for mapping lower-order brain functions such as vision, hearing, sensation, and movement, as well as higher-order brain functions, such as language, memory, and attention. Each function is typically mapped using a 3 - 5 minutes scan, which means that mapping multiple functional domains with tbfMRI would be exceedingly time consuming. While this may be acceptable in neuroscientific settings, where volunteers might tolerate longer scanning times and can be sufficiently trained for task performance, it is not usually tolerable for clinical patients whose task performance can degrade with longer scan times due to increasing discomfort, distraction or fatigue (Bennett & Miller, 2013; Hausman et al., 2022; Morrison et al., 2016). In addition, some patients may not be able to perform a task at all either due to cognitive impairment, lack of understanding, language or educational difficulties, very young or old age, or requiring sedation. All these issues are further compounded by the wide variation of functional mapping results due to specific task differences (Niskanen et al., 2012; Unadkat et al., 2019), and task-related head motion (Kochiyama et al., 2005).

In contrast, resting-state fMRI (rsfMRI) (Biswal et al., 1995), which measures the spontaneous fluctuation of BOLD signal and its correlation between different brain regions as functional connectivity, requires no task performance and a single scan of 5 - 7 minutes can typically be used to map multiple lower and higher-order resting-state networks (RSNs). RsfMRI can also be acquired with sedation, or while watching a video that can soothe very young patients who cannot hold still without an audio-visual stimulus (Pur et al., 2021). This makes it an attractive alternative to tbfMRI and assessing its applicability in a clinical setting is the focus of considerable debate and research. Multiple studies have shown good concordance between rsfMRI compared to intraoperative mapping for different functional domains (Cochereau et al., 2016; Lu et al., 2017; Qiu et al., 2014; Rosazza et al., 2014) . However, it suffers a number of limitations which may reduce its adoption in clinical neuroradiology. The absence of a task makes the interpretation of functional mapping based on rsfMRI functional connectivity more challenging. For example, a tbfMRI scan with a finger-tapping task shows the so-called functionally-eloquent areas of hand representation in the primary sensory-motor cortices, whereas rsfMRI shows the resting-state network (RSN) of sensory-motor (SM) function without directly differentiating eloquent and non-eloquent parts. There is also a lack of consensus on analysis methods, and a relative lack of clinical guidelines (Fox & Greicius, 2010; Gonzalez-Castillo et al., 2021; Lee et al., 2016; O’Connor & Zeffiro, 2019). Furthermore, while there are multiple data analysis tools for expert neuroscientists (Ashburner et al., 2006; Cox, 1996; Jenkinson et al., 2012), there are only a few user-friendly analysis packages (Hsu et al., 2018; Leuthardt et al., 2018; Whitfield-Gabrieli & Nieto-Castanon, 2012; Zacà et al., 2018) available for clinical research, and none provided by any major commercial health-tech vendors for rsfMRI. In contrast there is a wide variety of such clinically approved tools for tbfMRI data analysis.

Both task-based and rsfMRI data are subject to the confounding effect of head motion, EPI distortion and susceptibility effects. One recent development, multi-echo time (mTE) – fMRI aims to reduce these effects at the acquisition stage, and therefore has potential utility in the clinic.

MTE-fMRI acquires T2*-weighted images with at least 3 echo-times (TE) thus increasing signal-to-noise ratio (SNR) and providing a relative resilience to mild motion artifacts, which are independent of TE in contrast to the neuronal BOLD signal. In addition, it can partially recover BOLD signal in areas that are typically obscured by EPI distortion and susceptibility artifacts.

To the best of our knowledge, the relative accuracy of tbfMRI, sTE- and mTE-rsfMRI has not been validated against intraoperative functional mapping with DES in the same patient sample.

## Aim of the study

In this study we compared mapping results for the sensory-motor functions of the hands and feet from tbfMRI, sTE-rsfMRI at 2mm and 3mm voxel size, and mTE-rsfMRI to intraoperative functional mapping with DES using fully-automated methods. We quantitatively compared the three fMRI methods in a sample of 16 neurosurgical patients who underwent preoperative MRI-based and intraoperative DES-based functional mapping to investigate the suitability of the rsfMRI results for presurgical planning. We hypothesized that there would be no significant differences in the proximity of functional maps and DES coordinates among the four fMRI methods.

## Methodology

### Research questions

This study attempted to provide an in-depth understanding of the interplay between DES response, fMRI methods, fMRI tasks and their influence on proximity and agreement between fMRI activity and DES coordinates. To do so, we investigated the following research questions:

First **(RQ1)**, “How do the different fMRI methods compare in terms of raw distance measures between fMRI maps and DES coordinates?”, then **(RQ2)** “How do fMRI methods compare on receiver-operating characteristic (ROC) curves?”. Third, **(RQ3)** “How do fMRI methods compare in terms of binary agreement and disagreement with DES at different distance cutoffs? “, and lastly, **(RQ4)** “Are there significant differences between fMRI methods when accounting for the inter-subject repeated measures?”.

### Participants

We recruited 79 surgery-naïve patients who were referred to our department for presurgical fMRI and DTI between 01/2019 and 01/2021, 16 patients underwent presurgical tbfMRI and intraoperative mapping with matching cortical DES for the hands and/or feet. All participating patients, and/or their legal guardians signed written informed consent before participation, in accordance with the declaration of Helsinki. Local ethics committee approval was acquired (UZ/KU Leuven, Leuven, Belgium, study number S61759). Participating patients were excluded if they had undergone previous therapeutic brain surgery, had brain implants, e.g., deep brain stimulation electrodes, ventriculoperitoneal shunts, etc., or had absolute contraindications to MRI scanning. Summarized demographics and pathology information can be found in **Table 1** and further detailed in **S.table 1**.

**Table 1:** Summarized patient demographics and pathology information.

| Age and Gender | Lesion |  |  |
| --- | --- | --- | --- |
|  | Type | Laterality | Cerebral lobar distribution |
| Age range = 9 - 73 years<br>10 males<br>6 females<br>Median age = 39.5 years<br>Interquartile range = 28 years | 14 neoplasms:<br>13 gliomas (HGG)<br>1 meningioma<br>2 focal cortical dysplasia | 7 right sided<br>9 left sided | 2 fronto-parietal<br>1 fronto-temporal<br>8 frontal<br>2 parietal<br>1 parieto-occipital<br>1 Temporo-fronto-parietal<br>1 Fronto-temporo-parieto-occipital (multifocal lesion) |
| HGG = High grade glioma (World Health Organization grade > 2) |  |  |  |

### MRI acquisition

Two 3-Tesla whole-body MRI scanners were used for multimodal presurgical scanning (Ingenia -Elition, and Achieva DStream, Philips Medical Systems, Best, The Netherlands). Both MRI scanners were equipped with 32-channel phased-array receive head coils. The acquisition parameters for the 3D T1-weighted images, T2- and T2 fluid attenuation inversion recovery (FLAIR) images were previously described (Radwan et al., 2021). **Table 2** lists the acquisition parameters BOLD tbfMRI (single-echo), sTE-rsfMRI and mTE-rsfMRI data. The first patient (PT001) differed from the rest for sTE-rsfMRI, which was acquired with TR = 950 ms, multi-band = 8, voxel size = 2x2x2 mm, while PT002 had 250 volumes for both rsfMRI acquisitions.

**Table 2:** MRI acquisition parameters.

| Acquisition parameters/fMRI methods | tbfMRI | sTE-rsfMRI | mTE-rsfMRI |
| --- | --- | --- | --- |
| Acquisition plane - Pulse sequence | Axial - 2D gradient-echo EPI |  |  |
| TR/TE ms : FA ° | 1500/33 : 80 | 900/33 : 65 | 1150/8 - 33 - 58 : 75 |
| Voxel size mm<sup>3</sup> | 1.8x1.8x3.2 | 2x2x2.2 | 3x3x3 |
| Acquisition matrix | 112x112x44 | 112x112x66 | 80x80x48 |
| In-plane SENSE/Multiband SENSE | 2.1/3 | 1.2/6 | 1.9/4 |
| Pixel BW | 2162 | 2044 | 2253 |
| PE – Fat shit directions | AP – P |  |  |
| Number of volumes | 120-160 | 500 | 400 |
| MRI = magnetic resonance imaging, tbfMRI = task-based functional MRI, sTE-rsfMRI = single-echo resting-state fMRI, mTE-rsfMRI = multi-echo rsfMRI, EPI = echo planar imaging, TR = repetition time, TE = echo time, ms = milliseconds, FA = flip angle, SENSE = SENSitivity encoding in plane parallel imaging acceleration, BW = bandwidth, PE = phase encoding, AP = anteroposterior, P = posterior, DES = direct electrical stimulation |  |  |  |

### Functional MRI

A symmetrical block design consisting of 3 - 4 pairs of 30-second blocks of task performance versus neutral observation as a control condition was used for tbfMRI scanning. All tasks were explained and exercised before scanning. Sensory-motor tasks involved finger-tapping or fist-clenching for the hands, and movement of the toes. Patients were instructed to engage in mind-wandering for rsfMRI while a black screen was shown. This was substituted with watching a video or movie if the patient was unable to remain still without a visual stimulus. Visual stimuli were synchronized with the scanner and displayed using Presentation (Neurobehavioral Systems, NC, USA) via a projector system to a flat in-bore plastic projector screen or via an MRI-compatible display. In both cases, the visual stimuli, e.g., start, move, and stop, rest, were visible to the patient via a mirror mounted on top of the head coil and MRI-compatible corrective glasses were used if needed. Tb-fMRI and sTE-rsfMRI were acquired for all patients, while mTE-rsfMRI was not acquired for 2 patients (PT006 and PT010).

### Intraoperative brain mapping

Wake-up neurosurgery and cortical intraoperative DES were performed for the 16 patients included in this study. Intraoperative frameless neuronavigation (Curve, BrainLab, Munich, Germany) was employed in all cases. Cortical DES used the OSIRIS neurostimulator (Inomed Medizintechnik GmbH, Germany), and a bipolar fork stimulator with 5mm inter-electrode spacing (Inomed) for cortical mapping. Stimulation parameters followed the protocols described by Duffau et al (Duffau et al., 1999) (60Hz) in anaesthetized patients and the low frequency protocol described by Zangaladze et al. (Zangaladze et al., 2008) (5Hz) in awake patients. **Table 3** lists the DES tests done, the number of resulting positive and negative DES coordinates, effects of positive DES responses, and fMRI tasks of interest for each patient.

**Table 3:** DES intraoperative brain mapping protocol details.

| Coded patient names | Awake surgery | MEP/SSEP | Stimulation range in mA | Cortical / subcortical threshold |
| --- | --- | --- | --- | --- |
| PT001 | yes | no | 4 - 20 mA | 20 mA / - |
| PT002 | no | yes | 4 - 14 mA | 6 mA / 4 mA |
| PT003 | yes | no | 4 - 6 mA | 4 mA |
| PT004 | yes | no | 4 - 20 mA | - / 5 mA |
| PT005 | yes | no | 4 - 20 mA | 16 mA / 5mA |
| PT006 | yes | no | 4 - 20 mA | 16 mA / 10 mA |
| PT007 | yes | no | 4 - 20 mA | 20 mA / 2 mA |
| PT008 | yes | no | 2 - 20 mA | 8 mA / - |
| PT009 | yes | no | 4 - 20 mA | - |
| PT010 | yes | no | 4 - 20 mA | - |
| PT011 | yes | no | 4 - 20 mA | 12 mA / - |
| PT012 | yes | no | 4 - 20 mA | 12 mA / 5 mA |
| PT013 | yes | no | 4 - 20 mA | 20 mA / 5 mA |
| PT014 | yes | no | 4 - 20 mA | 20 mA / 10 mA |
| PT015 | yes | no | 4 - 20 mA | - / 10 mA |
| PT016 | yes | no | 4 - 15 mA | 4 mA / 4 mA |

MRI = magnetic resonance imaging, tbfMRI = task-based functional MRI, sTE-rsfMRI = single-echo resting-state fMRI, mTE-rsfMRI = multi-echo rsfMRI, TFE = turbo field echo, EPI = echo planar imaging, TR = repetition time, TE = echo time, ms = milliseconds, FA = flip angle, SENSE = SENSitivity encoding in plane parallel imaging acceleration, CS = compressed SENSE, BW = bandwidth, PE = phase encoding, AP = anteroposterior, F = feet, P = posterior, DES = direct electrical stimulation, MEP/SSEP = motor and somatosensory evoked potentials, mA = milliamperere

### Data analysis

Figure 1 shows a schematic representation of the data preprocessing and analysis workflow. Data conversion, lesion segmentation with ITK-snap v3.8.0 (Yushkevich et al., 2019), lesion inpainting with KU Leuven Virtual Brain Grafting v0.52 (KUL_VBG) (Radwan et al., 2021) (https://github.com/KUL-Radneuron/KUL_VBG), and parcellation were previously described (Radwan et al., 2023). Briefly, we constructed 5 mm radius spheres centered around each DES coordinate, then calculated minimum Euclidean distances between the center of gravity (COG) of each DES sphere and every voxel in the corresponding functional map. Additionally, we calculated similarity measures using dice similarity coefficient (DSC) and Jaccard index (JI) between corresponding functional maps from the 4 BOLD fMRI methods. All acquired images were converted to the brain imaging data structure format (BIDS) (Gorgolewski et al., 2016) format using the KULeuven Neuroimaging suite (KUL_NIS) (*KULeuven Neuro Imaging Suite (KUL_NIS)*, 2018/2022) (https://github.com/treanus/KUL_NIS) and dcm2bids (*Dcm2bids*, 2016/2022).

**Figure 1.**
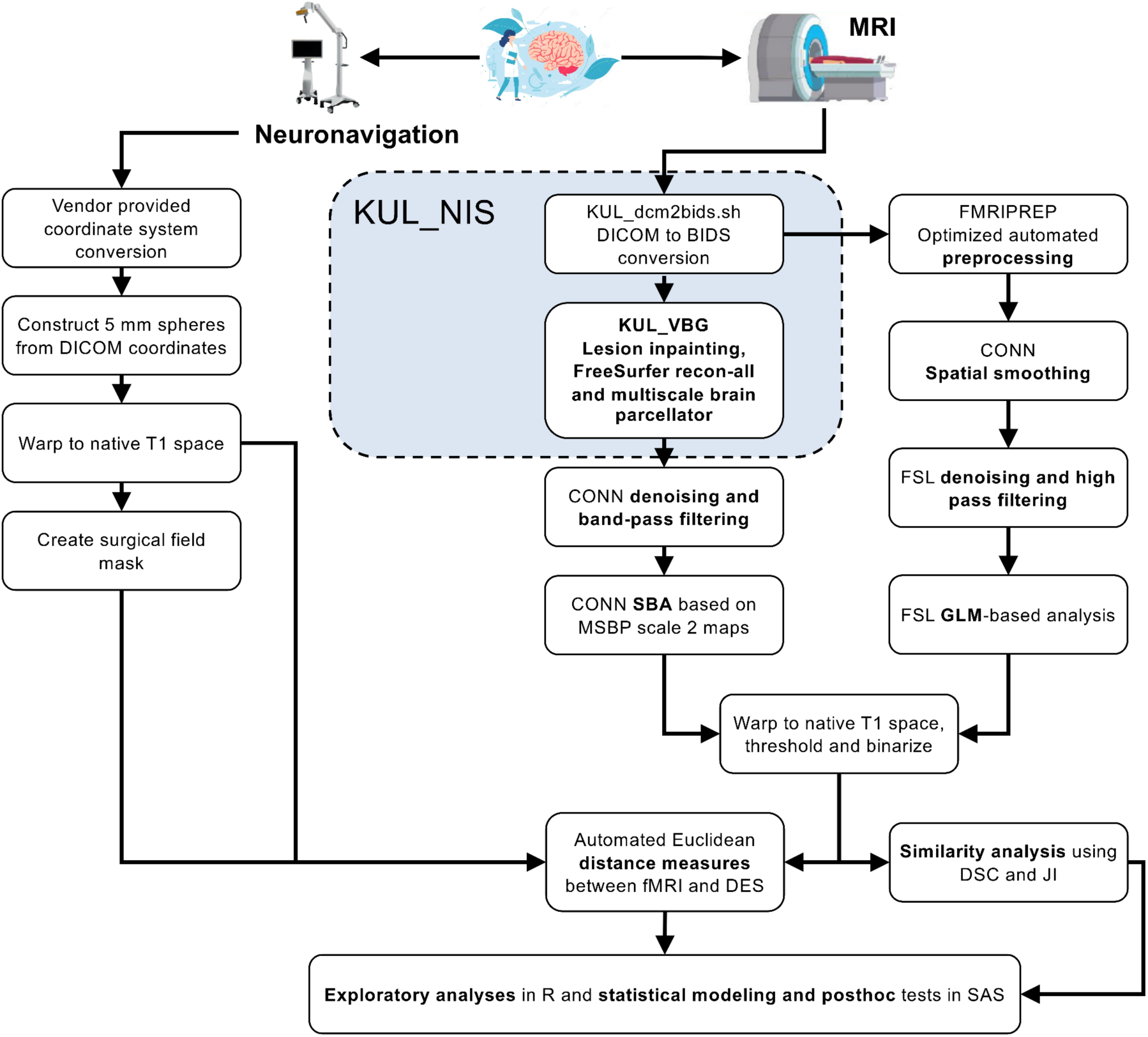
Schematic representation of the data preprocessing and analysis workflow used to compare different fMRI results to intraoperative mapping outcome. MRI = magnetic resonance imaging, KUL_NIS = KU Leuven neuroimaging suite, BIDS = brain imaging data structure, KUL_VBG = KU Leuven virtual brain grafting, CONN = functional connectivity toolbox, SBA = seed-based analysis, GLM = general linear model, DSC = dice similarity coefficient, JI = Jaccard index

### BOLD fMRI data analysis

All BOLD fMRI data were preprocessed using fmriprep v20.2.6 (Esteban et al., 2019), which combines methods from different software packages in an optimized preprocessing pipeline. This corrected for slice-timing, motion artifacts, EPI-included image distortion, calculated various covariates for denoising, and applied inter-modality warping between the BOLD and anatomical images, as well as normalization to the asymmetrical MNI152 nonlinear 2009 template with 2 mm isotropic voxels (MNI152NLin2009cAsym_res-2). The middle echo images (TE = 33 ms) of the mTE-rsfMRI series were also used separately as the sTE-rsfMRI 3mm method, which perfectly matches the mTE-rsfMRI data in acquisition parameters, and patient state. Fmriprep was used for combining the different echoes of mTE-rsfMRI data into a single time-series (Posse et al., 1999). All preprocessed BOLD fMRI data were imported in the functional connectivity analysis toolbox (CONN) (Whitfield-Gabrieli & Nieto-Castanon, 2012) and smoothed with a 3D gaussian kernel of 6 mm full-width at half-maximum. ANTs v2.3.0 (Avants et al., 2011; Tustison et al., 2021) was used to warp resulting maps to native T1 space.

### Task-based fMRI processing

Spatially-smoothed tbfMRI images generated by CONN were brain extracted, then denoised by regressing out the normalized framewise displacement using fsl_glm FSL v6.0 (Jenkinson et al., 2012), and high-pass filtered using fslmaths with σ = 20 TRs. Results were analyzed using a general linear model (GLM) in fsl_glm with default settings other than specifying a double gamma HRF convolution. Output Z-score maps were then warped back and resampled to native T1 space for further analysis.

### Resting-state fMRI processing

Preprocessed rsfMRI (sTE and mTE) were denoised and analyzed using the seed-to-voxel approach in CONN (Whitfield-Gabrieli & Nieto-Castanon, 2012) and default covariates. Default band-pass filtering (0.001 – 0.01 Hz) was applied after nuisance regression. The MSBP (Tourbier et al., 2020) scale-2 parcellation maps were propagated into subject-specific binary grey matter masks using ANTs then imported to CONN to define the seeds for functional connectivity analysis. Seed-to-voxel maps for hands, and feet were derived from second-level GLM analysis by calculating the average group-level connectivity maps. We used bilateral precentral part 3 labels as seeds for the hands, and bilateral paracentral labels for the feet. The resulting subject-specific beta maps from CONN’s second-level GLM were used for further analysis.

### Thresholding of tbfMRI and rsfMRI results

Results of tbfMRI and rsfMRI processing were constrained by a smoothed subject-specific grey-matter tissue mask derived from the normalized T1-weighted images, then warped back to native T1 space with antsApplyTransforms. K-means thresholding was applied with 100 thresholds using ANTS ThresholdImage, which rescaled the positive intensities in the maps to similar values (1 - 101). This was followed by applying a minimum threshold to the Kmeans images, which was defined as:

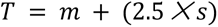

Where T is the minimum threshold, m is the mean value of nonzero voxels, and *s* is the standard deviation of non-zero voxels. Resulting thresholded fMRI maps were binarized for further analysis.

### DES coordinates processing, distance and volume measures

Distance measures were limited to the surgical field using subject-specific binary voxel masks to minimize the contribution of potential spurious fMRI results from outside the surgical field. This was done by summing all DES spheres, then binarizing the result and applying a 3D morphological dilation filter with σ = 15 mm using ANTs ImageMath, then masking the outcome with the binary brain mask. Minimum Euclidean distances were calculated between all DES coordinates represented by the COG of each DES sphere and all voxels of the corresponding fMRI map within the surgical field mask in Python 3.8 using nibabel v3.2.2 (Brett et al., 2022), numpy v1.22.3 (C. R. Harris et al., 2020), and scipy v1.4.1 (Virtanen et al., 2020). The whole fMRI map was used if no valid fMRI voxels were found within the surgical field mask. Continuous distance measures were rounded to their closest integers (in mm) because increments smaller than a single voxel (1mm) were not considered meaningful.

### fMRI similarity measures

Dice similarity coefficient (DSC), and Jaccard index (JI) were calculated between functional maps for the hands and feet generated with tbfMRI and each rsfMRI method for the same subject, as well as between the rsfMRI methods using ANTS (Tustison et al., 2021) LabelOverlapMeasures. Differences in similarity measures between fMRI methods were explored using descriptive statistics.

### Statistical testing

#### Exploratory analysis

Comparing different fMRI methods when a ground truth is present may be achieved with techniques typically employed to compare screening tests, such as confusion matrices. However, due to the small sample size, unequal number of DES samples and functional maps per patient, we first plotted the distance measures (Patil, 2021) without imposing any cutoff for agreement **(RQ1)**. Then, to account for the unequal repetition of DES coordinates per subject, the distance measures were averaged for nDES and pDES separately, so that each patient had at most 1 pDES and 1 nDES measurement for hands and/or feet. These averaged measures were then used to create ROC (Robin et al., 2011) curves to explore differences in sensitivity, specificity, and to estimate distance cutoffs for further analysis of the unaveraged data **(RQ2)**. Different distance cutoffs were estimated based on the local maxima of the averaged tbfMRI data and used to evaluate the rsfMRI methods. DeLong tests were used for direct pairwise comparison of the ROC curves.

Next, we explored differences in binary agreement and disagreement at the ROC determined distance cutoffs **(RQ3)**, excluding subjects with missing modalities (PT006 and PT010). Positive results were represented by fMRI-DES pairs with a distance less than the cutoff, true-positives if involving pDES, and false-positives if involving nDES coordinates. Negative results were represented by fMRI-DES pairs with a distance above the cutoff, true-negatives if involving nDES and false-negatives if involving pDES. Lastly, two-part linear models were used to compare all fMRI-DES distance measures between fMRI methods while accounting for the unequal intrasubject DES repetition **(RQ4)** at the different distance cutoffs determined on the tbfMRI ROC.

#### Two-part linear modeling

Distance data thresholded at the three cutoffs determined by ROC of all pooled data were used for a two-part linear mixed model and posthoc testing to compare fMRI methods while accounting for the unequal number of DES coordinates between different subjects. The thresholded distances data were a semicontinuous variable with excess zeros and an extremely right-skewed distribution, violating assumptions of normality. Therefore, and given the within-subjects nesting of repeated distance measures, we opted for a two-part model for longitudinal data (Farewell et al., 2017; Tooze et al., 2002) **(RQ3)**. The model was estimated with the %MIXCORR macro provided by Tooze, Gunwald and Jones, 2002(Tooze et al., 2002) and PROC NLMIXED in SAS studio v9.4 (SAS Institute, Cary, NC, USA).

The first part (A) predicted the probability of overlap (distance=0), and the second part (B) predicted the distances between nonoverlapping (distance>0) fMRI-DES coordinate pairs. Distances were the dependent variable and DES response, and fMRI method (tbfMRI, sTE-rsfMRI 2mm, sTE-rsfMRI 3mm, and mTE-rsfMRI) were used as predictors in both parts of the model, and adaptive Hochberg’s (Hochberg & Benjamini, 1990) family-wise error-rate (FWE) correction was used to control for type(I) error in posthoc testing. No covariates were used in this analysis due to the small sample size; further detail can be found in supplementary material.

## Results

### Lesion segmentation

Volumetric lesion voxel-masks included perilesional edema in case of neoplasms. The median lesion volume was 44.70 ml, minimum = 1.20, maximum = 124.78, and IQR = 46.61 ml. Figure 2 shows the voxel-wise probabilistic distribution of lesions in this sample of patients over the whole brain in the MNI152 space.

**Figure 2.**
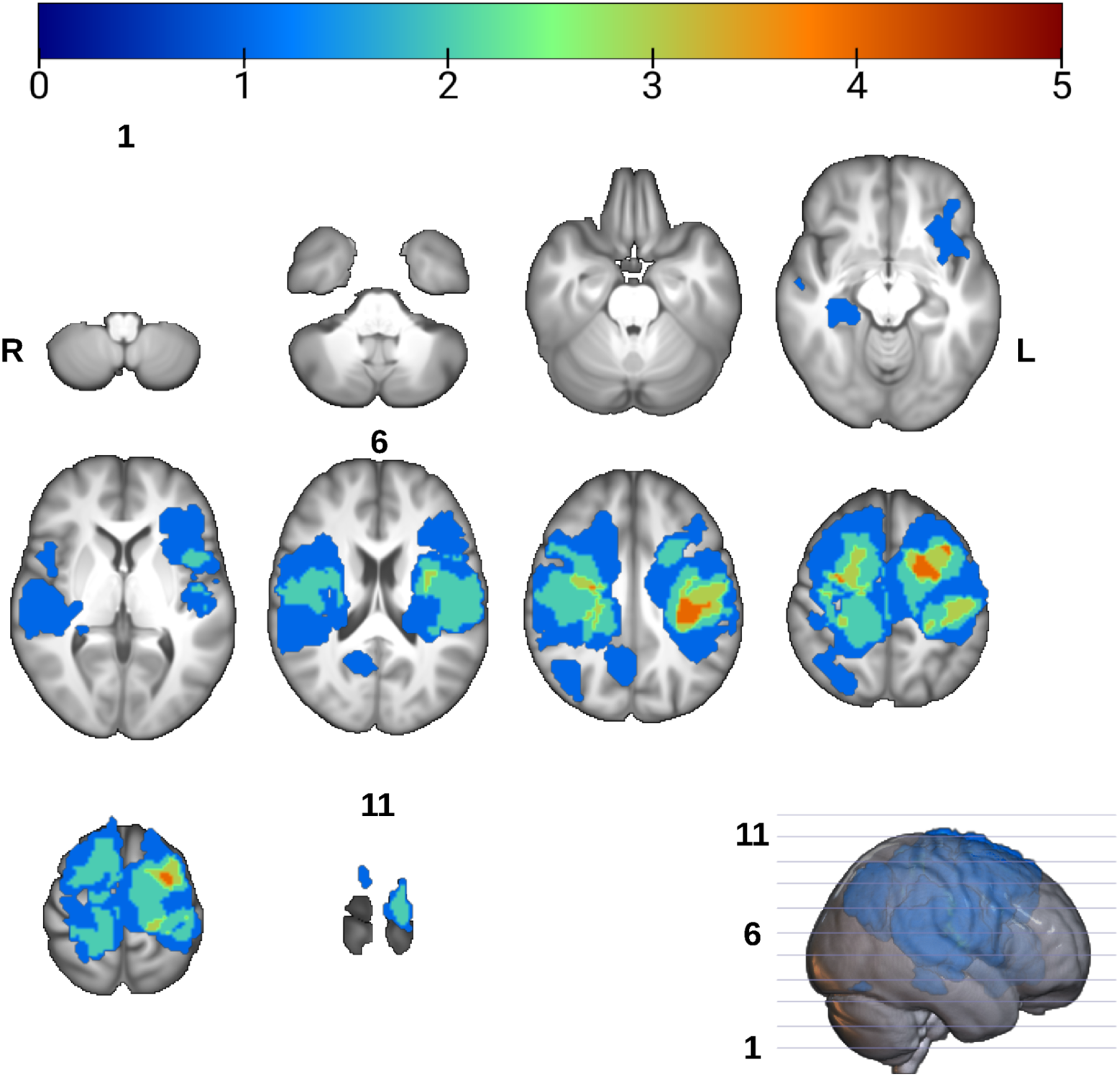
Spatial distribution of lesions from all patients overlaid onto the UK biobank T1 template brain in standard Montreal neurological institute (MNI) space. Overlay voxel intensities correspond to the sum of lesion masks occupying it. R = right, L = Left, slice numbers are indicated for the first, middle and last slices

### BOLD fMRI mapping

All fMRI data was successfully processed resulting in 22 tbfMRI maps, 22 sTE-rsfMRI maps, and 20 mTE-rsfMRIs as patients PT006, and PT010 did not undergo the mTE-rsfMRI scan. Hands were mapped for 16 patients using tbfMRI and sTE-rsfMRI, and for 14 patients using mTE-rsfMRI, and feet were mapped for 6 patients using the 4 methods. All resulting maps were included for further analysis regardless of the amount of motion during scanning. Figures 3 **and 4** show all resulting fMRI maps for the hands and feet tasks, respectively.

**Figure 3.**
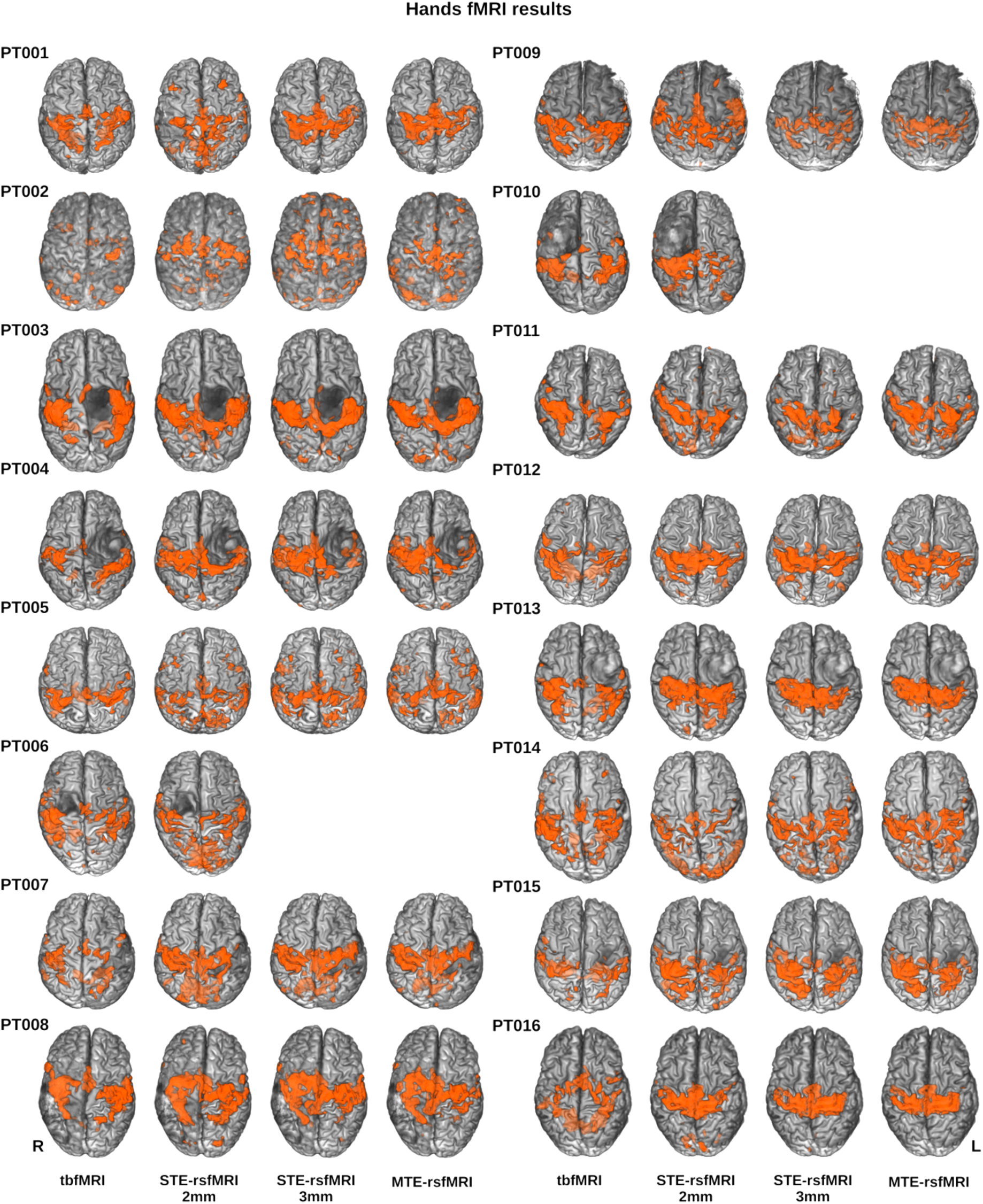
Results for hands fMRI mapping using all methods overlaid in orange on surface rendered T1 images for each patient in superior view, empty cells indicate a missing multi-echo rsfMRI scan, tbfMRI = task-based fMRI, sTE-rsfMRI = single-echo resting-state fMRI, mTE-rsfMRI = multi-echo rsfMRI

**Figure 4.**
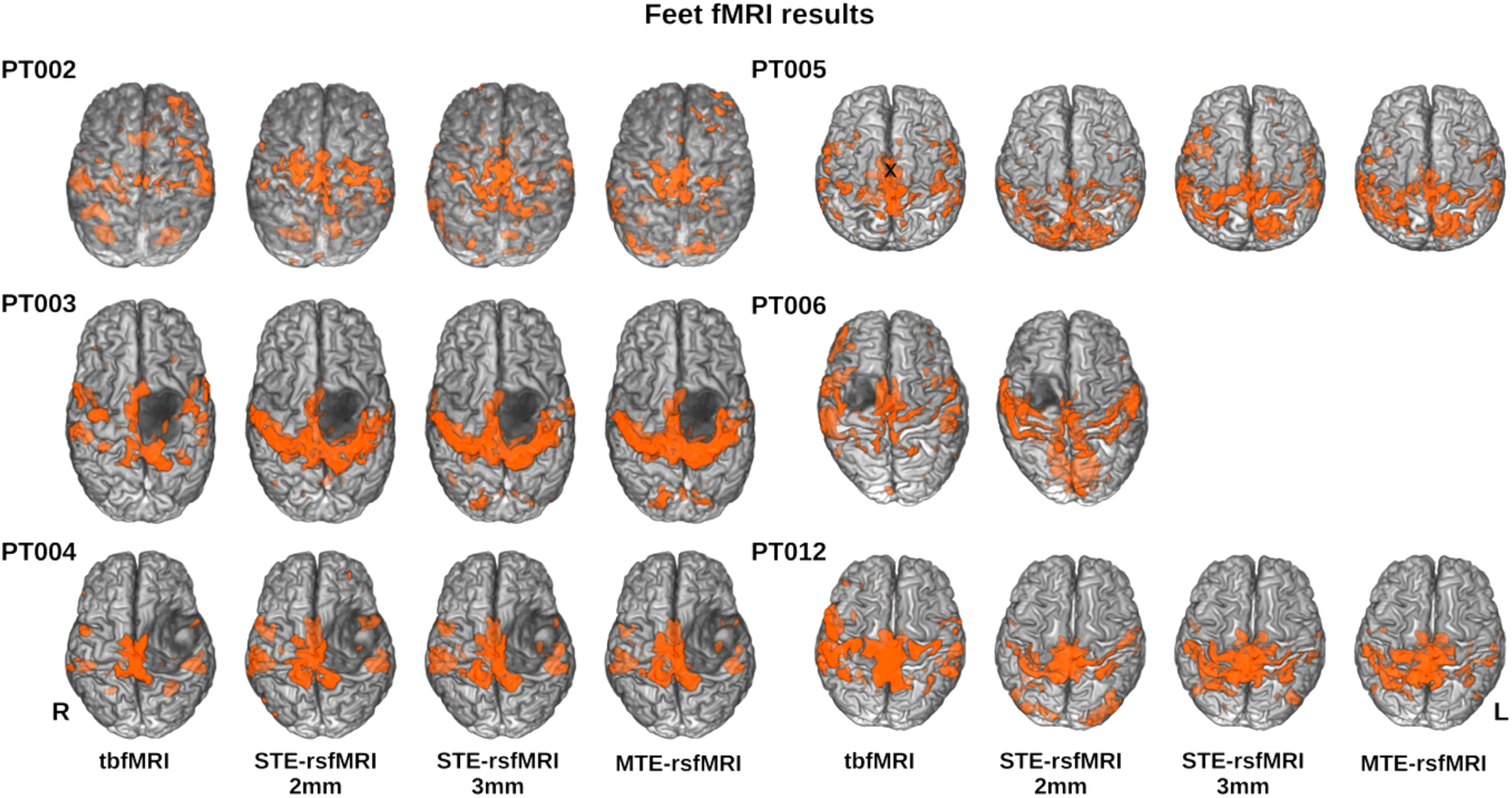
Results for feet fMRI mapping using all methods overlaid in orange on surface rendered T1 images for each patient in superior view, tbfMRI = task-based fMRI, sTE-rsfMRI = single-echo resting-state fMRI, mTE-rsfMRI = multi-echo rsfMRI

### Intraoperative mapping and distance measures

DES mapping resulted in a total of 23 positive DES (pDES) and 88 negative DES (nDES) coordinates. Figure 5 shows images from exemplar patients demonstrating DES spheres and example functional mapping results included in the surgical field mask. Further details on results of intraoperative DES mapping can be found in **S.table 2**. Functional maps were paired with the relevant pDES spheres and all nDES spheres, which resulted in 512 distance measures in total with 138 measures for tbfMRI and sTE-rsfMRI 2mm and 118 for sTE-rsfMRI 3mm and mTE-rsfMRI. See **S.figure 1** for plots of distance measures per DES response.

**Figure 5.**
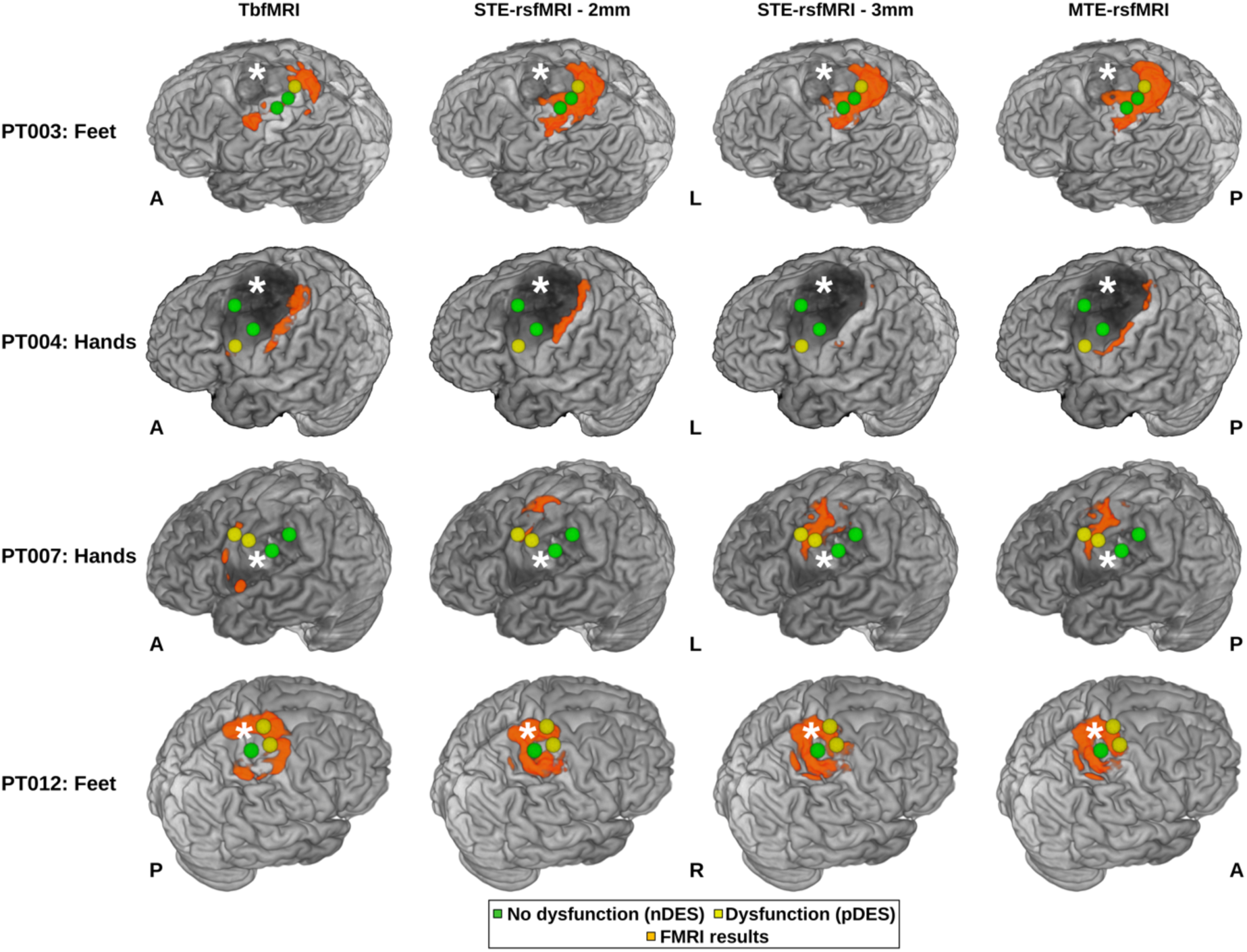
DES spheres and fMRI mapping results for hands and feet using with the 3 methods overlaid on top of surface-rendered T1 images. White asterisks indicate site of pathology, PT = patient, nDES = negative direct electrical stimulation, pDES = positive DES, tbfMRI = task-based fMRI, sTE-rsfMRI = single-echo resting-state fMRI, mTE-rsfMRI = multi-echo rsfMRI, A = anterior, P = posterior, L = left, R = right, S = superior, I = inferior

### Similarity analysis

In terms of inter-method intrasubject similarity regardless of task, DSC and JI scores showed modest similarity between tbfMRIs and the three rsfMRI methods, with sTE-rsfMRI 2mm (DSC/JI median = 0.203/0.113, IQR = 0.159/0.103) scoring lower than mTE-rsfMRI (DSC/JI median = 0.260/0.150, IQR = 0.136/0.087) and sTE-rsfMRI 3mm (DSC/JI median = 0.284/0.166, IQR = 0.169/0.109). Higher similarity measures were found between sTE-rsfMRI 2mm and mTE (DSC/JI median = 0.536/0.367, IQR = 0.341/0.288), sTE-rsfMRI 3mm and mTE-rsfMRI (DSC/JI median = 0.557/0.728, IQR = 0.248/0.229), and sTE-rsfMRI 2mm and 3mm (DSC/JI median = 0.546/0.376. IQR = 0.345 /0.301). DSC and JI were found to be higher for the hands (DSC/JI median = 0.333/0.200, IQR = 0.358/0.326) than for feet (DSC/JI median = 0.269 /0.164, IQR = 0.306/0.348), see **S.table 3a and 3b** for more details.

### Statistical testing

#### Exploratory analysis

Excluding PT006 and PT010, who did not undergo mTE-rsfMRI, only minor differences were found between fMRI methods for unthresholded distance measures **(RQ1)**, see **table 4** and **S.figure 1**. ROC curves local maxima determined three cutoffs of 3.7, 6.5, and 10.1 (integers: 4, 7, and 10) mm for the averaged tbfMRI data, **(RQ2)** see **table 5** and figure 6 for detailed results, and **S.figure 2** for ROCs generated from unaveraged distances. Briefly for the averaged data, tbfMRI had the highest sensitivity, specificity, and area-under the curve, AUC = 92.1%, followed by sTE-rsfMRI 2mm, which had comparable sensitivity, mildly lower specificity and AUC = 88.2%. The reduced voxel size seemed to induce a reduction in accuracy, as sTE-rsfMRI 3mm showed AUC = 82.1%, and mTE-rsfMRI scored the lowest, AUC = 79.3%. All methods showed lower sensitivity and higher specificity at the lowest cutoff, and higher sensitivity and lower specificity at higher cutoffs. DeLong tests showed no significant differences between any of the fMRI methods, see **S.table 4**.

**Figure 6.**
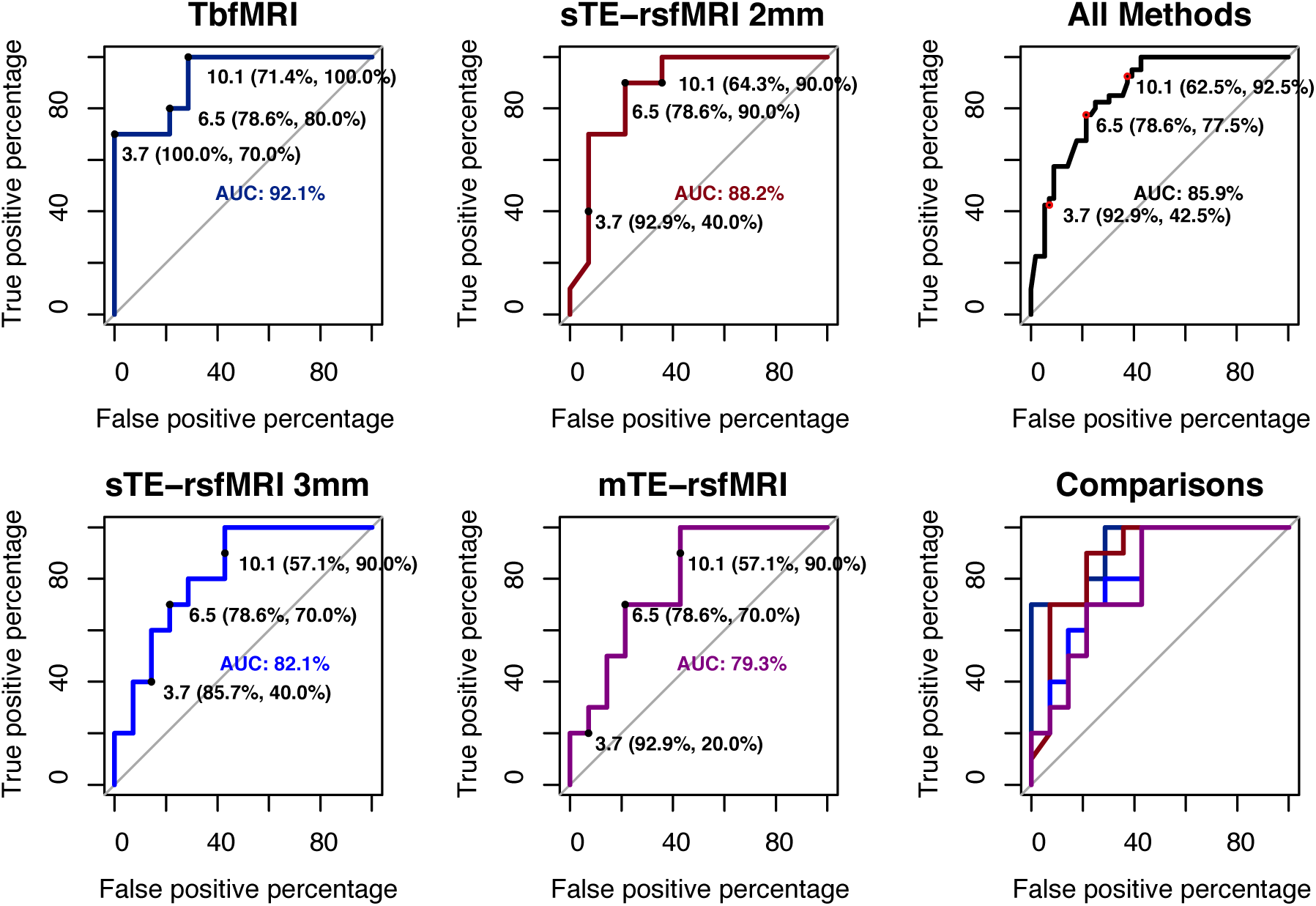
ROCs plots and distance cutoffs per fMRI method, the corresponding sensitivity and specificity values are shown as well as the area under the curve (AUC) value per method, tbfMRI = task-based fMRI, sTE-rsfMRI = single-echo resting-state fMRI, mTE-rsfMRI = multi-echo rsfMRI

**Table 4:** Descriptive statistics for distance measures per DES response, function and fMRI method.

| Function | DES response | fMRI method | Mean | StDev | Median | IQR | Var | Range | N |
| --- | --- | --- | --- | --- | --- | --- | --- | --- | --- |
| Hands | pDES | TbfMRI | 4.176 | 3.225 | 4 | 6 | 10.404 | 10 | 17 |
|  |  | sTE-rsfMRI 2 mm | 4.647 | 3.426 | 5 | 4 | 11.742 | 12 | 17 |
|  |  | sTE-rsfMRI 3 mm | 4.933 | 4.0789 | 4 | 4 | 16.638 | 14 | 15 |
|  |  | mTE-rsfMRI | 6.066 | 3.6344 | 6 | 4.5 | 13.209 | 13 | 15 |
|  | nDES | TbfMRI | 14.534 | 10.085 | 12 | 14 | 101.722 | 45 | 88 |
|  |  | sTE-rsfMRI 2 mm | 15.886 | 10.655 | 14 | 14.25 | 113.550 | 46 | 88 |
|  |  | sTE-rsfMRI 3 mm | 15.389 | 10.917 | 13 | 16.75 | 119.183 | 38 | 76 |
|  |  | mTE-rsfMRI | 15.618 | 10.527 | 13 | 17.25 | 110.825 | 42 | 76 |
| Feet | pDES | TbfMRI | 2.166 | 1.471 | 2.5 | 1.75 | 2.166 | 4 | 6 |
|  |  | sTE-rsfMRI 2 mm | 2.666 | 2.160 | 2.5 | 2.5 | 4.666 | 6 | 6 |
|  |  | sTE-rsfMRI 3 mm | 3.6 | 3.209 | 2 | 2 | 10.3 | 8 | 5 |
|  |  | mTE-rsfMRI | 3.4 | 3.209 | 2 | 1 | 10.3 | 8 | 5 |
|  | nDES | TbfMRI | 13.925 | 12.034 | 10 | 12 | 144.840 | 50 | 27 |
|  |  | sTE-rsfMRI2 mm | 13.851 | 12.024 | 9 | 16.5 | 144.592 | 37 | 27 |
|  |  | sTE-rsfMRI3 mm | 14.181 | 13.022 | 10.5 | 24 | 169.584 | 36 | 22 |
|  |  | mTE-rsfMRI | 15.409 | 13.730 | 10 | 24.75 | 188.538 | 40 | 22 |

DES = direct electrical stimulation, pDES = positive DES, nDES = negative DES, fMRI = functional magnetic resonance imaging, StDev = standard deviation, IQR = interquartile range, Var = variance, N = number, tbfMRI = task-based fMRI, sTE-rsfMRI = single echo resting-state fMRI, mTE-rsfMRI = multi-echo resting-state fMRI

**Table 5:** ROC derived accuracy measures at different cutoffs.

| ROC accuracy measures at different distance cutoffs 4/7/10mm for averaged and raw distance measures |  |  |  |  |
| --- | --- | --- | --- | --- |
| Method | Sensitivity % |  | Specificity % |  |
|  | Averaged | Raw | Averaged | Raw |
| **TbfMRI** | 70/80/100 | 56.5/78.3/100 | 100/78.6/71.4 | 88.7/73/56.5 |
| **STE-rsfMRI 2mm** | 40/90/90 | 47.8/82.6/91.3 | 92.9/78.6/64.3 | 88.7/75.7/57.4 |
| **STE-rsfMRI 3mm** | 40/90/90 | 45/80/90 | 85.7/78.6/57.1 | 84.7/70.4/54.1 |
| **MTE-rsfMRI** | 20/70/90 | 35/70/90 | 92.9/78.6/57.1 | 88.8/72.4/58.2 |

TbfMRI = task-based fMRI, STE-rsfMRI = single echo resting-state fMRI, MTE-rsfMRI = multi-echo resting-state fMRI

Plots for binary agreement and disagreement rates showed only minor differences between fMRI methods at all ROC-determined distance cutoffs **(RQ3)**, see **S.figures 3 and 4**.

#### Two-part linear modeling

Below we describe the results of statistical modeling of predicted probability of agreement between fMRI-DES pairs where the distance measured was below the cutoff and predicted distance in case of disagreement (distance > cutoff) **(RQ4)**. Results of the logistic regression part (model A) are followed by the results of the lognormal part (model B) for the 3 ROC-determined distance cutoffs.

Differences between the 4 fMRI methods were not significant at any of the ROC-determined distance cutoffs, indicating that there were no significant differences in predicted probability of overlap and predicted distances between tbfMRI, sTE-rsfMRI (2mm and 3mm), and mTE-rsfMRI. Differences between DES response types were significant (p< 0.001) in both parts of the model at the 3 cutoffs, meaning that pDES coordinates were associated with significantly higher probabilities of overlap and shorter distances to fMRI activity regardless of the method used. No significant differences were found between hands and feet, indicating that the two domains behave similarly in both parts of the model. The results are listed in **table 6**. and illustrated visually in Figure 7.

**Figure 7.**
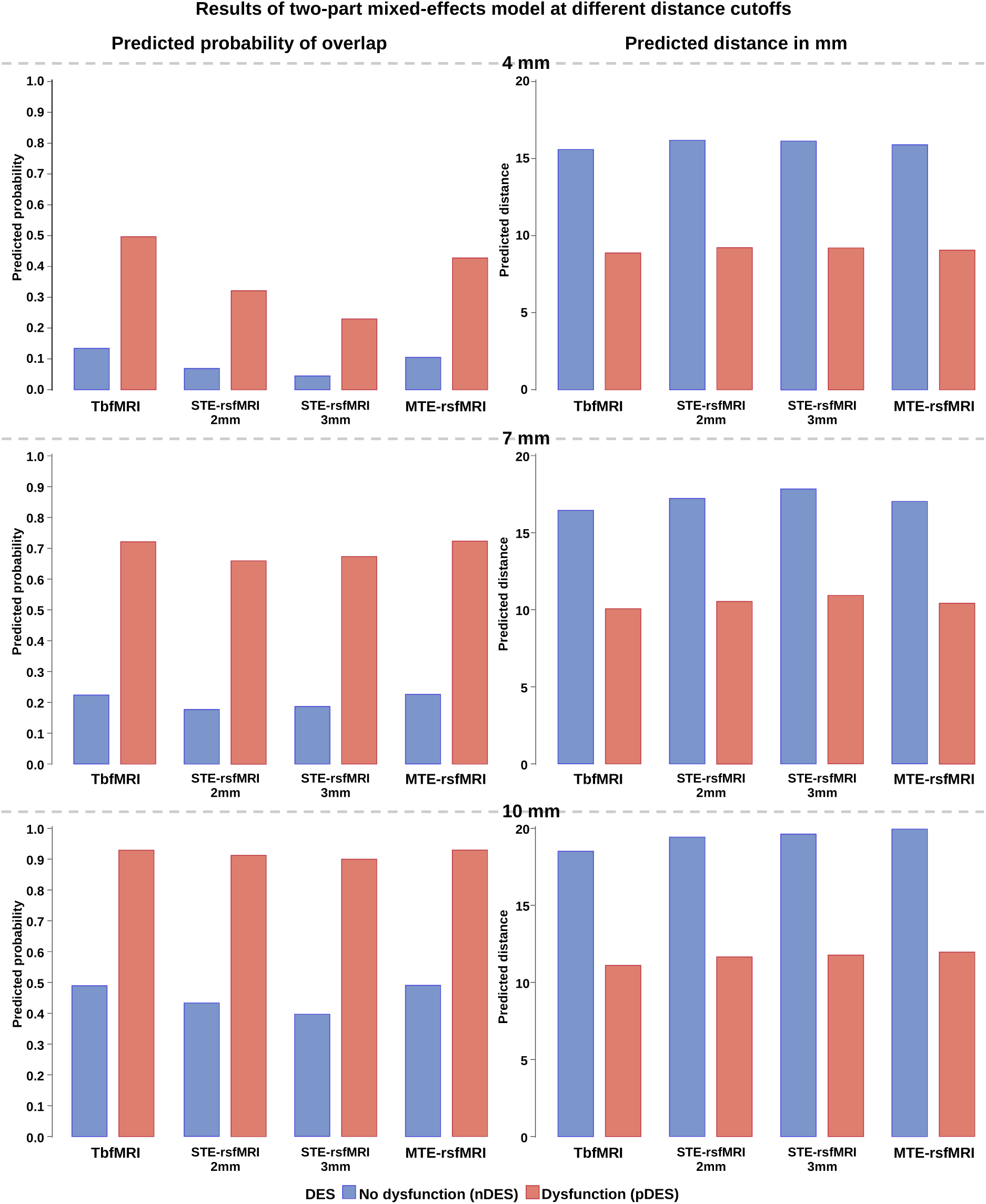
Results for predicted probability of overlap and predicted distances for nonoverlapping DES spheres averaged over fMRI tasks at different distance cutoffs for defining agreement, nDES = negative direct electrical stimulation, pDES = positive DES, tbfMRI = task-based fMRI, sTE-rsfMRI = single-echo resting-state fMRI, mTE-rsfMRI = multi-echo rsfMRI

**Table 6:** Summarized results of the estimated two-part models for predicting probability of overlap and distance between fMRI and DES.

| Distance threshold for agreement |  |  | 4 mm |  | 7 mm |  | 10 mm |  |
| --- | --- | --- | --- | --- | --- | --- | --- | --- |
| Parameter | Model | DF | t Value | Pr > \|t\| | t Value | Pr > \|t\| | t Value | Pr > \|t\| |
| tbfMRI v STE-rsfMRI 2mm | A | 14 | -1.69 | 0.226 | -0.78 | 0.980 | -0.70 | 0.988 |
| tbfMRI v STE-rsfMRI 3mm | A | 14 | -0.64 | 0.531 | 0.03 | 0.980 | -0.02 | 0.988 |
| tbfMRI v mTE-rsfMRI | A | 14 | -2.49 | 0.052 | -0.59 | 0.980 | -1.02 | 0.988 |
| STE-rsfMRI 2mm v 3mm | A | 14 | 1.00 | 0.531 | 0.77 | 0.980 | 0.65 | 0.988 |
| STE-rsfMRI 3mm v MTE | A | 14 | -1.88 | 0.163 | -0.60 | 0.98 | -0.98 | 0.988 |
| tbfMRI v STE-rsfMRI 2mm | B | 14 | -0.52 | 0.963 | -0.67 | 0.878 | -0.72 | 0.881 |
| tbfMRI v STE-rsfMRI 3mm | B | 14 | -0.26 | 0.963 | -0.47 | 0.878 | -1.04 | 0.881 |
| tbfMRI v mTE-rsfMRI | B | 14 | -0.45 | 0.963 | -1.11 | 0.878 | -0.84 | 0.881 |
| STE-rsfMRI 2mm v 3mm | B | 14 | 0.23 | 0.963 | 0.16 | 0.878 | -0.38 | 0.881 |
| STE-rsfMRI 3mm v MTE | B | 14 | -0.18 | 0.963 | -0.62 | 0.878 | 0.22 | 0.881 |

Model A = logistic regression part evaluating probability of overlap/agreement at distance cutoff, Model B = lognormal regression part evaluating distance measures in case of nonoverlap, DF = degrees of freedom, Pr > |t| = probability, TIV = total intracranial volume, sTE-rsfMRI = single-echo resting-state fMRI, mTE-rsfMRI = multi-echo rsfMRI

## Discussion

The main aim of this study was to compare the accuracy of tbfMRI and rsfMRI using DES results as the ground truth. We did so first **(RQ1)** by comparing fMRI methods for the unthresholded distance measures. This showed minor differences between the fMRI methods for mapping hands and/or feet when compared to pDES and nDES. ROCs were then used to compare measures of accuracy, sensitivity and specificity between the fMRI modalities **(RQ2)**. As tbfMRI represents the routine standard of practice for clinical fMRI mapping, its averaged distance values were used to estimate the distance cutoffs. This analysis showed that (a) tbfMRI was the most accurate while sTE-rsfMRI 2mm scored slightly worse. (b) The change in acquisition parameters, and patient state between the sTE-rsfMRI 2mm scan and the mTE-rsfMRI scan results in an apparent reduction in accuracy relative to DES. (c) The distance cutoff of 7mm appeared to maximize both sensitivity and specificity on averaged data ROCs. Next, we compared fMRI methods for binary agreement measures at the 3 cutoffs **(RQ3)**, which also showed only minor differences between fMRI methods when compared to pDES and nDES. Similarly, the two-part mixed-effects linear model showed only minor and non-significant differences between the fMRI methods at the three distance cutoffs **(RQ4)**.

Given that mTE-rsfMRI would be expected to perform at least as well as sTE-rsfMRI, its worse performance on ROC curves was unexpected. While this was also apparent on plotting in **S**.**Figure 1**, and figure 6, it was not reflected in the results of the two-part mixed-effects models at any of the distance cutoffs, as would be expected for a small difference.

The results of the similarity analysis with DSC and JI were in line with previous studies, which reported rather low agreement between tbfMRI and rsfMRI despite good concordance with DES (Rosazza et al., 2014). In this case, the low overlap may be partially attributed to differences in acquisition parameters, which undoubtedly played a role. Similarity scores can be expected to improve with matched acquisition parameters and optimized seed-selection for SBA, as evidenced by the higher DSC and JI (0.557 and 0.728) between sTE-rsfMRI 3mm and mTE-rsfMRI than between sTE-rsfMRI 2mm and sTE-rsfMRI 3 mm (0.536 and 0.367). This may be expected as tbfMRI and rsfMRI in fact measure different aspects of neural activity and therefore could be expected to give mildly similar but not identical results (Dierker et al., 2017). Yet, despite the low similarity, we found no significant differences in accuracy between tbfMRI and sTE-rsfMRI, indicating that both methods can be used for accurate SMN mapping.

This study adds to the growing body of evidence that presurgical functional brain mapping with rsfMRI is feasible with comparable accuracy to tbfMRI. In contrast to previous studies, here we used fully-automated data analysis methods that accounted for the presence of pathology such as KUL_VBG (Radwan et al., 2021) for lesion inpainting and minimizing subsequent errors in resulting parcellation maps, as well as advanced statistical modeling. In addition, we generated hands and feet specific seed-based rsfMRI maps by relying on the finer-grained parcellation maps from MSBP (Tourbier et al., 2020), which to the best of our knowledge, was previously done in only a few studies either using manual delineation (Rosazza et al., 2014; Schneider FC, 2015) or by repeating ICA within the SMN mask (Sohn et al., 2012). Only a small number of studies have included different fMRI methods and DES results as a gold standard in their analyses (Cui et al., 2022; Mitchell et al., 2013; Roland et al., 2019; Rosazza et al., 2014; Vakamudi et al., 2020; Zacà et al., 2018; Zhang et al., 2009), none of which used automated parcellation-based SBA for mapping hands and feet from rsfMRI, or included mTE-rsfMRI.

While the currently dominant paradigm in neurosurgical practice prioritizes mapping the eloquent sensory-motor, and/or language areas for preservation during surgery, recent evidence has shown the importance of mapping and preservation of functional networks that are generally thought of as non-eloquent. Higher-order RSNs such as the ventral and dorsal attention networks, salience network, default mode network and executive control networks if injured may be associated with reduced patient independence, and increased morbidity and mortality postoperatively (Dadario et al., 2021).

RsfMRI may serve as a clinically viable and more accessible alternative to tbfMRI. Considering the complexity of the tasks associated with mapping functions using tbfMRI, the significance of rsfMRI is underscored by its task-free nature, its ability to provide maps that are modestly similar to those of tbfMRI, and its comparable distance measures to DES coordinates in this sample of clinical patients. Despite recent studies indicating that these RSNs can also be mapped from tbfMRI data by regressing out task-related signal changes (R. J. Harris et al., 2014; Pareto et al., 2018) there is an expectation of higher reliability of RSNs from rsfMRI compared to tbfMRI data analyzed with this approach. This expectation arises because a single rsfMRI scan typically acquires more time-points (volumes) than a single tbfMRI scan, and a higher number of volumes has been correlated with increased reliability of mapped RSNs (Birn et al., 2013; White et al., 2014). It is important to note that while rsfMRI may be considered a viable alternative in case tbfMRI is not possible, if the patient is cooperative, tolerant to scanning and if sufficient scan time is available, a combined acquisition of tbfMRI and rsfMRI remains a more data rich approach. Additionally, analysis of task-independent signal change in tbfMRI data can still be expected to offer valuable information.

## Study limitations

In contrast to recent studies (Dierker et al., 2017; Ngo et al., 2022; Niu et al., 2021; Parker Jones et al., 2017), we did not employ machine learning or deep learning methods for predicting tbfMRI from the rsfMRI data. While such studies show highly encouraging results, the majority are not easily accessible for clinical validation. Furthermore, these techniques typically require larger curated datasets for training and testing, and the pretrained models, if provided, might not translate easily to data from different scanners. Among the limitations of this study are the small sample size, non-standardization of sensory-motor mapping tasks between fMRI and DES, patients sample heterogeneity in terms of age and pathology, as well as not including information on patient symptoms and postoperative follow up. Lastly, mTE-fMRI was only evaluated only as a resting-state technique, and only 3 TEs were acquired, while a higher number of TEs can be expected to improve mapping outcomes.

## Conclusion

By using DES as the ground truth to compare measures of accuracy between tbfMRI, sTE-rsfMRI and mTE-rsfMRI, we have demonstrated that automated parcellation driven SBA sTE-rsfMRI can be used for presurgical brain mapping of sensory-motor representation of the hands and feet. Further investigation in a larger sample, preferably with denser sampling during invasive mapping, is necessary to further explore the lower accuracy of mTE-rsfMRI and sTE-rsfMRI acquired at 3mm, as well as the generalizability of these findings to different sites and different functional networks.

## Data Availability

All data produced in the present study are available after anonymisation upon reasonable request to the authors within the limitations imposed by GDPR

## Supplementary information

### Two-part linear model details

The first part (A) used a logit-link for binary response (distance=0 vs. distance >0) and a generalized linear mixed model to predict probability of nonoverlap (distance > 0), and the second part (B) used a log-normal linear mixed model for the distance measures (distance >0) between nonoverlapping fMRI-DES coordinate pairs.

The thresholded distance measures were used as the dependent variable and DES response type (positive and negative), fMRI methods (tbfMRI, sTE-rsfMRI 2mm, sTE-rsfMRI 3mm, and mTE-rsfMRI 2mm) were used as predictors in both parts of the model.

For ease of interpretation, we discuss and plot the probability of overlap (distance = 0) for the first part (A), and log distances (B) from the second part are back-transformed to distance in mm.

Results were interpreted in the following context: Predicted probability of overlap (distance < cutoff) with pDES coordinates was analogous to true-positive rate. Predicted probability of overlap with nDES coordinates was analogous to false-positive rate. Predicted distances to non-overlapping (distance > cutoff) nDES coordinates were analogous to true-negative rate and predicted distances to non-overlapping pDES coordinates were analogous to false-negative rate.

In this analysis, missing data was represented and handled by assigning ’NAN’ (Not a Number) values. The decision to use ’NAN’ ensured that the missing data did not influence the statistical calculations or skew the results. The analysis was then carried out on the remaining dataset, excluding the ’NAN’ values.

---

**Supplementary figure 1.**
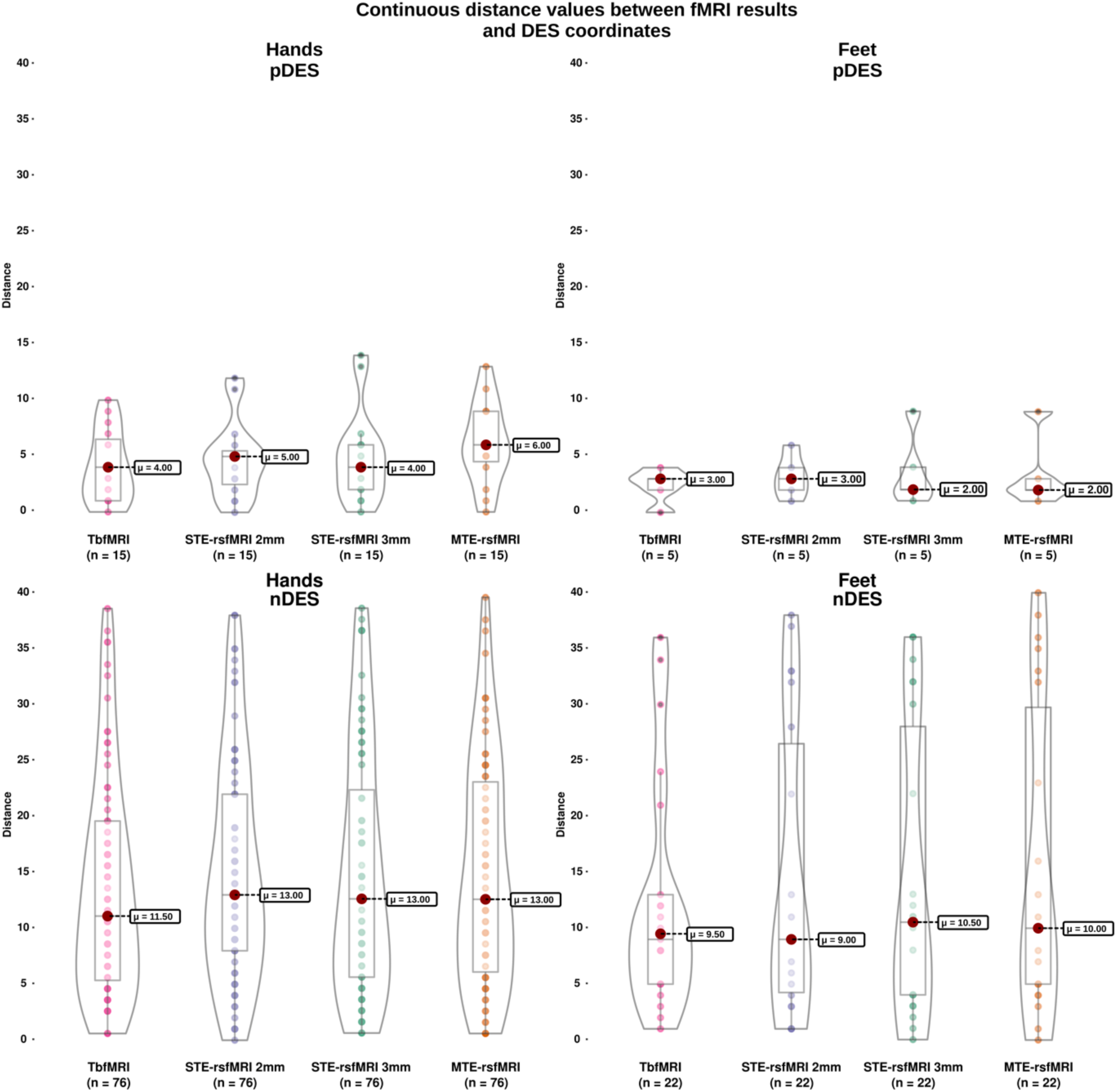
Distance measures for fMRI and DES responses shown in box and violin plots. Results for the hands are shown on the left, for the feet on the right, for pDES on top, and for nDES on the bottom, tbfMRI = task-based fMRI, sTE-rsfMRI = single-echo resting-state fMRI, mTE-rsfMRI = multi-echo rsfMRI

**Supplementary figure 2.**
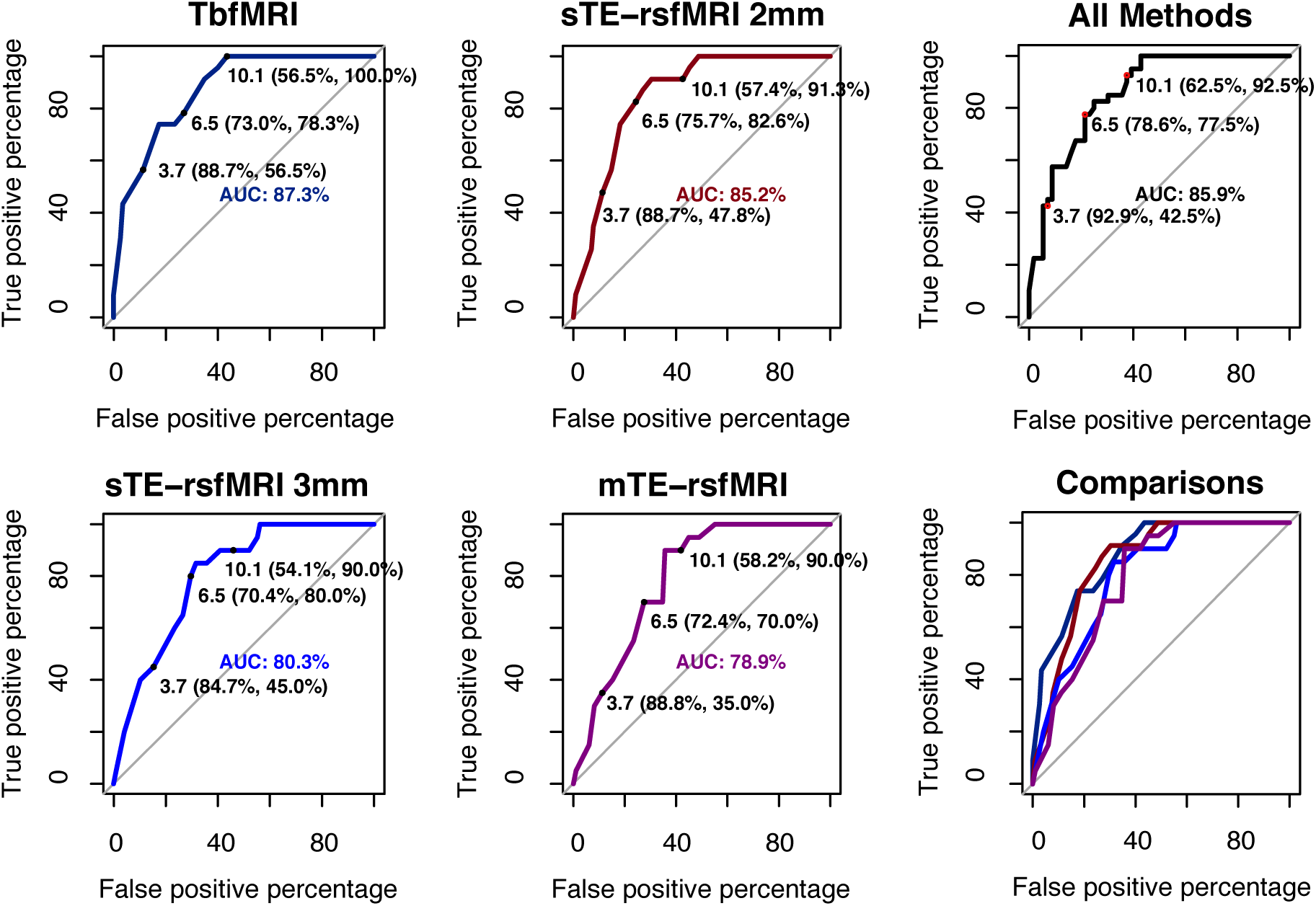
ROCs of raw distance measures without intra-subject averaging.

**Supplementary figure 3.**
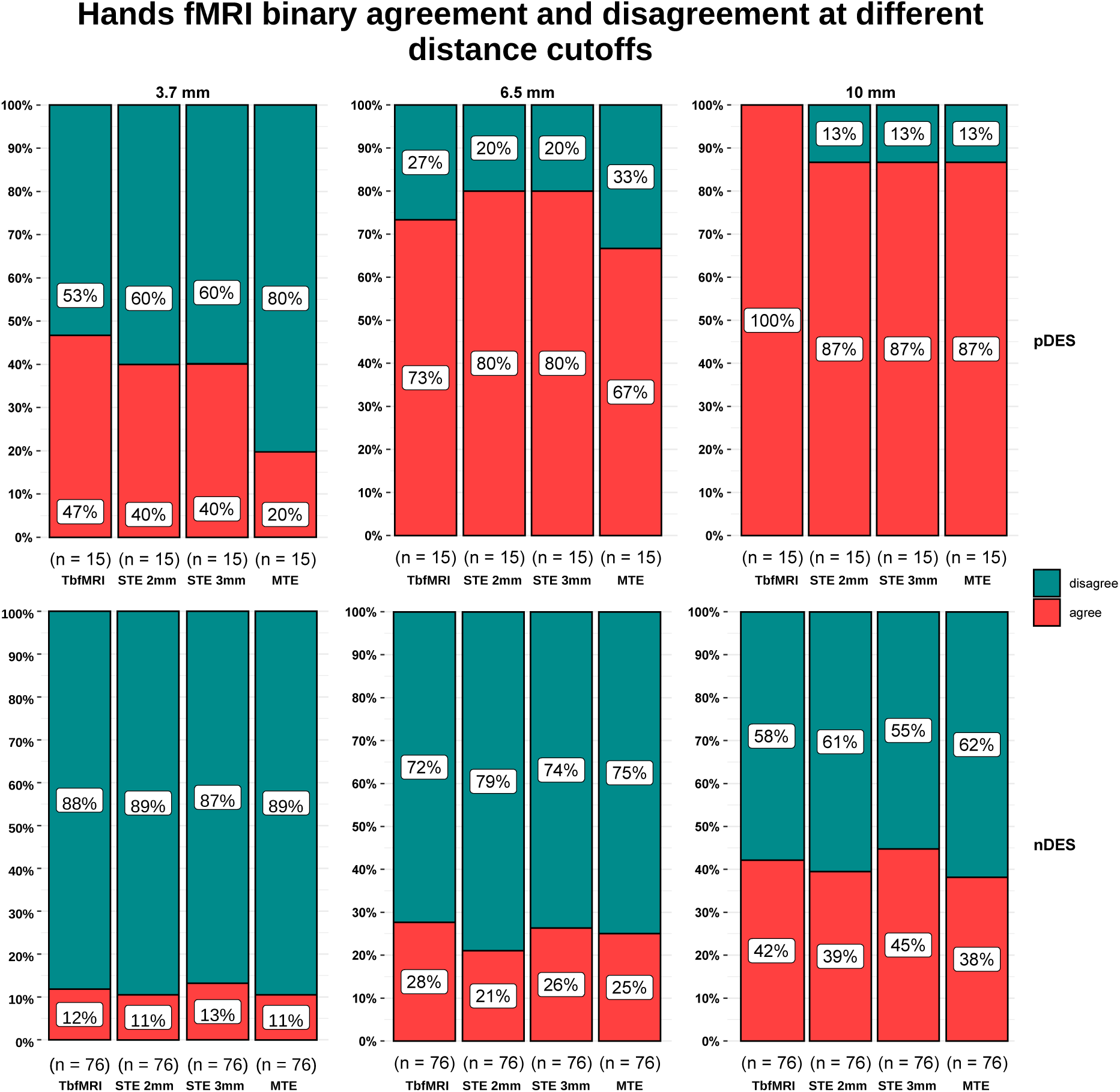
Stacked column plots depicting hands fMRI percent of agreement versus disagreement with different fMRI methods and DES at ROC-determined distance cutoffs, tbfMRI = task-based fMRI, sTE = single-echo resting-state fMRI, mTE = multi-echo rsfMRI

**Supplementary Figure 4.**
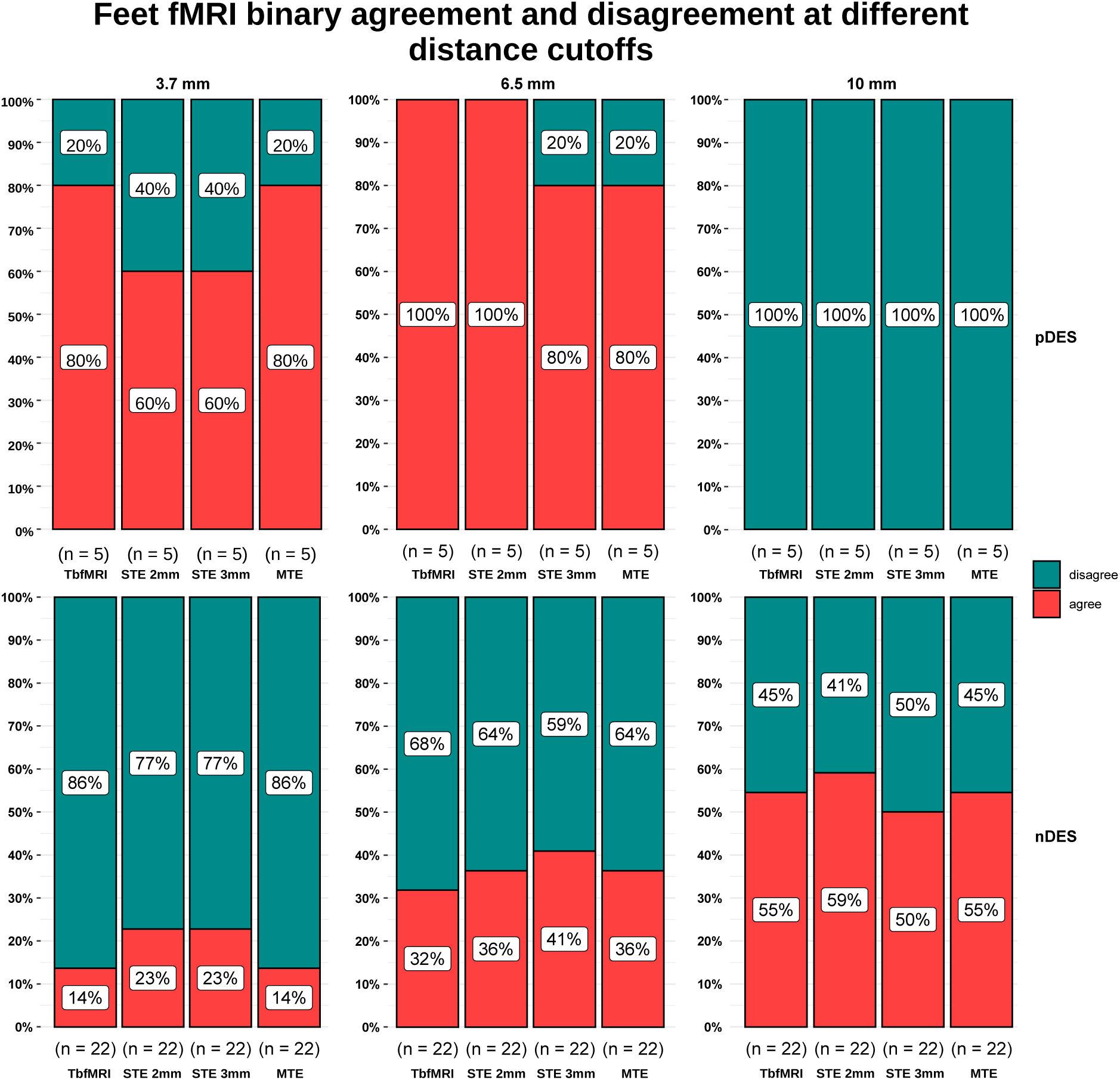
Stacked column plots depicting feet fMRI percent of agreement versus disagreement with different fMRI methods and DES at ROC-determined distance cutoffs, tbfMRI = task-based fMRI, sTE-rsfMRI = single-echo resting-state fMRI, mTE-rsfMRI = multi-echo rsfMRI

**Supplementary table 1:** Patient demographics, pathology results, lesion lobe, side and volume.

| Coded patient names | Age at scan | Gender | Lesion type | WHO grade | Pathology report | Lesion side and lobe | Lesion volume (ml) |
| --- | --- | --- | --- | --- | --- | --- | --- |
| PT001 | 61 – 65 | M | Glioma | IV | Glioblastoma | R / Fr-Pa | 66.809 |
| PT002 | 6 – 10 | F | FCD |  | Type I | L / Fr | 1.200 |
| PT003 | 36 – 40 | M | Meningioma | I | Transitional type meningioma | L / Fr | 54.079 |
| PT004 | 31 – 35 | F | Glioma | II | Oligodendroglioma | L / Fr-Pa | 124.775 |
| PT005 | 71 – 75 | M | Glioma | IV | Glioblastoma | R / Pa-Oc | 32.382 |
| PT006 | 66 – 70 | F | Glioma | IV | Glioblastoma | R / Fr | 26.779 |
| PT007 | 31 – 35 | M | Glioma | IV | Glioblastoma | L / Te-Fr-Pa | 88.917 |
| PT008 | 41 – 45 | M | Glioma | II | Multifocal astrocytoma | R / Fr-Te-Pa-Oc | 123.293 |
| PT009 | 61 – 65 | F | Glioma | II | Oligodendroglioma | L / Fr | 53.331 |
| PT010 | 31 – 35 | M | Glioma | III | Oligodendroglioma | R / Fr | 92.154 |
| PT011 | 31 – 35 | F | Glioma | II | Oligodendroglioma | L / Pa | 22.553 |
| PT012 | 61 – 65 | M | Glioma | IV | Glioblastoma | R / Pa | 42.517 |
| PT013 | 31 – 35 | F | Glioma | III | Oligodendroglioma | L / Fr | 46.887 |
| PT014 | 56 – 60 | M | Glioma | III | Astrocytoma | L / Fr-Te | 40.208 |
| PT015 | 56 – 60 | M | Glioma | II | Oligodendroglioma | L / Fr | 11.531 |
| PT016 | 11 - 15 | M | FCD |  | Type IIB | R / Fr | 4.806 |

PT = patient, M = male, F = female, FCD = focal cortical dysplasia, WHO = world health organization, R = right, L = left, ml = milliliters, Fr-Pa = frontoparietal, Fr = frontal, Pa-Oc = parieto-occipital, Te-Fr-Pa = Temporo-occipito-parietal, Fr-Te-Pa-Oc = Fronto-temporo-parieto-occipital, Pa = parietal, Fr-Te = fronto-temporal

**Supplementary table 2:** Direct electrical stimulation (DES) mapping and fMRI details.

| Coded patient names | Test done | pDES/nDES | DES positive effect | N. of fMRI methods / tasks |
| --- | --- | --- | --- | --- |
| PT001 | Active motor and language mapping | 0/6 | none | 4 / Hands |
| PT002 | Active motor mapping | 4/6 | Motor response right upper leg, wrist, foot, and hand | 4 / Right hand and foot |
| PT003 | Active motor mapping | 3/3 | Motor response right upper leg, foot, and hand | 4 / Hands and feet |
| PT004 | Active motor and language mapping | 1/7 | Motor response right hand, and lower arm | 4 / Hands and feet |
| PT005 | Active motor mapping | 2/4 | Interference with finger tapping, and motor response left hand | 4 / Hands and feet |
| PT006 | Active motor mapping | 2/6 | Sensory-motor response left leg | 2 / Hands and feet |
| PT007 | Active motor and language mapping | 0/5 | Motor response right hand | 4 / Hands |
| PT008 | Active motor mapping | 0/6 | none | 4 / Hands |
| PT009 | Active motor mapping | 0/7 | none | 4 / Hands |
| PT010 | Active motor mapping | 1/6 | Motor response left hand and foot | 2 / Hands |
| PT011 | Active motor mapping | 1/6 | Motor response right hand | 4 / Hands |
| PT012 | Active motor mapping | 2/2 | Motor response left foot, leg, hand and arm | 4 / Hands and feet |
| PT013 | Active motor mapping | 2/8 | Motor response right hand | 4 / Hands |
| PT014 | Active motor mapping | 1/6 | Motor response right hand | 4 / Hands |
| PT015 | Active motor and language mapping | 1/4 | Motor response right arm | 4 / Hands |
| PT016 | Active motor mapping | 3/6 | Motor response left wrist and hand | 4 / Hands |

PT = patient, DES = direct electrical stimulation, tbMRI = task-based functional magnetic resonance imaging, sTE-rsfMRI = single-echo resting-state fMRI, mTE-rsfMRI = multi-echo rsfMRI, AF = arcuate fasciculus, CST = corticospinal tract, MEP/SSEP = motor and somatosensory evoked potential, B0 = non-diffusion weighted spin-echo EPI volume, 2 = tbfMRI and sTE-rsfMRI, 3 = tbfMRI, sTE-rsfMRI, and mTE-rsfMRI

**Supplementary table 3:** Summarized descriptive statistics for similarity measures per fMRI technique (3a) and per fMRI task (3b)

| **Supplementary table 3a:** Summarized descriptive statistics for similarity measures per fMRI technique |  |  |  |  |
| --- | --- | --- | --- | --- |
| **Methods/Measures** | **Metric** | **Max(min)** | **Mean(std)** | **Median(IQR)** |
| **sTE-rsfMRI 2mm v tbfMRI** | **DSC** | 0.516(0.061) | 0.235(0.122) | 0.203(0.159) |
|  | **JI** | 0.348(0.032) | 0.138(0.082) | 0.113(0.103) |
| **sTE-rsfMRI 3mm v tbfMRI** | **DSC** | 0.526(0.0277) | 0.254(0.139) | 0.284(0.169) |
|  | **JI** | 0.357(0.014) | 0.153(0.094) | 0.166(0.109) |
| **mTE-rsfMRI v tbfMRI** | **DSC** | 0.450(0.029) | 0.239(0.121) | 0.260(0.136) |
|  | **JI** | 0.290(0.015) | 0.140(0.078) | 0.150(0.087) |
| **sTE-rsfMRI 2mm v 3mm** | **DSC** | 0.730(0.045) | 0.471(0.221) | 0.546(0.345) |
|  | **JI** | 0.576(0.023) | 0.333(0.181) | 0.376(0.301) |
| **sTE-rsfMRI 2mm v mTE-rsfMRI** | **DSC** | 0.740(0.044) | 0.454(0.219) | 0.536(0.341) |
|  | **JI** | 0.588(0.022) | 0.318(0.180) | 0.367(0.288) |
| **sTE-rsfMRI 3mm v mTE-rsfMRI** | **DSC** | 0.744(0.099) | 0.525(0.190) | 0.557(0.248) |
|  | **JI** | 0.871(0.161) | 0.655(0.199) | 0.728(0.229) |
| **Supplementary table 3b:** Summarized descriptive statistics for similarity measures per fMRI task |  |  |  |  |
| **Tasks/Measures** | **Metric** | **Max(min)** | **Mean(std)** | **Median(IQR)** |
| **Hands** | **DSC** | 0.744(0.028) | 0.367(0.203) | 0.333(0.358) |
|  | **JI** | 0.851(0.014) | 0.291(0.228) | 0.200(0.326) |
| **Feet** | **DSC** | 0.740(0.053) | 0.338(0.226) | 0.269(0.306) |
|  | **JI** | 0.871(0.027) | 0.271(0.242) | 0.164(0.348) |
| sTE-rsfMRI = singe-echo resting-state fMRI, tbfMRI = task-based fMRI, mTE-rsfMRI = multi-echo rsfMRI, DSC = Dice similarity coefficient, JI = Jaccard index, max = maximum, min = minimum, stdev = standard deviation, IQR = interquartile range |  |  |  |  |

**Supplementary table 4:** Summarized results of DeLong pairwise tests comparing the ROC curves from averaged distance measures.

| Pairwise comparisons | Estimate1 | Estimate2 | Statistic | p.value | CI.low | CI.high |
| --- | --- | --- | --- | --- | --- | --- |
| TbFMRI v sTE-rsfMRI 2mm | 92.14 | 88.21 | 0.53 | 0.60 | -0.11 | 0.18 |
| TbFMRI v sTE-rsfMRI 3mm | 92.14 | 82.14 | 1.53 | 0.13 | -0.03 | 0.23 |
| TbFMRI v mTE-rsfMRI | 92.14 | 79.29 | 1.55 | 0.12 | -0.03 | 0.29 |
| sTE-rsfMRI 2mm v 3 mm | 88.21 | 82.14 | 1.12 | 0.26 | -0.05 | 0.17 |
| sTE-rsfMRI 2mm v mTE-rsfMRI | 88.21 | 79.29 | 1.50 | 0.13 | -0.03 | 0.21 |
| sTE-rsfMRI 3mm v mTE-rsfMRI | 82.14 | 79.29 | 0.57 | 0.57 | -0.07 | 0.13 |

ROC = receiver operating characteristic, CI = confidence interval, TbFMRI = task-based fMRI, sTE-rsfMRI = single-echo resting-state fMRI, mTE-rsfMRI = multi-echo resting-state fMRI

